# Predictors of intraoperative resectability among pancreatic cancer patients for curative-intent surgery: a systematic review with meta-analysis

**DOI:** 10.64898/2026.01.01.26343303

**Authors:** Vernon Chuabio, Rochelle Cion, Carl Lawrence Arenos

## Abstract

**Background:** Some pancreatic cancer patients for curative-intent surgery are found to actually have unresectable disease on laparotomy. This systematic review aimed to identify parameters that can potentially help predict intraoperative resectability status.

**Methods:** Literature search was done across PubMed, Scopus, Web of Science, and Cochrane Library to identify studies since 2011 assessing for independent association of preoperative parameters with intraoperative pancreatic cancer resectability. Parameters investigated by at least two studies were identified. For discordant findings, meta-analyses using the random effects model were done, incorporating studies that can be pooled. Pooled odds ratios and mean differences were obtained for parameters with binary and continuous values, respectively, with 95% confidence intervals. Subgroup analyses were done for significant heterogeneity.

**Results:** From initially 8,959 articles searched, 18 studies were included, with 13 parameters identified for synthesis. Elevated serum CA19-9 (>35-1000 U/mL) and larger tumor size (>22-40 mm) were shown to independently predict intraoperative unresectability in the most number of studies. For parameters subjected to meta-analyses, abdominal pain, weight loss, lower body mass index (BMI), lower serum transaminases, and preoperative borderline resectability were significantly associated with intraoperative unresectability, while age and sex were not. Subgroup analyses addressing heterogeneity revealed that tumor location in the pancreatic body/tail is significantly associated with actual unresectability whereas poorer performance status was not. Other parameters shown to have predictive value but could not be included in meta-analyses are serum CA125 and imaging-to-surgery time interval.

**Conclusion:** Synthesis of studies has shown that elevated serum CA19-9 and larger tumors are independently associated with intraoperative pancreatic cancer unresectability. Other predictive factors include abdominal pain, weight loss, lower BMI, lower serum transaminases, preoperative borderline resectability, and tumor location in the pancreatic body/tail. Staging laparoscopy or neoadjuvant therapy should be considered for potentially resectable pancreatic cancer patients with these attributes.

**What is already known on this topic:** Accurate identification of patients with actual resectable pancreatic cancer remains challenging despite resectability criteria. Some parameters beyond resectability criteria have been found to offer clues regarding actual resectability.

**What this study adds:** Synthesis of studies has determined which clinical, laboratory, and radiologic parameters outside of resectability criteria may predict intraoperative resectability among pancreatic cancer patients being considered for curative-intent surgery.

**How this study might affect research, practice, or policy:** The findings in this study may aid in deciding which pancreatic cancer patients should undergo staging laparoscopy first instead of directly attempting curative surgery. Research gaps have also been identified.

## Background

Surgical resection remains the best chance to have the best outcomes among pancreatic cancer patients.^1,2^ The decision to perform curative-intent surgery on pancreatic cancer patients is mainly determined by radiologic assessment of resectability, which includes evaluation of vascular involvement and distant metastasis.^3^ However, some patients subjected to attempted curative resection are found to have actual unresectable disease on surgical exploration, resulting in aborting the curative-intent procedure.^4^ An international consensus by the International Association of Pancreatology (IAP) has identified biological factors including serum CA19-9 level and performance status that may herald distant metastasis even among patients with potentially resectable pancreatic cancer.^5^ The National Comprehensive Cancer Network (NCCN) has suggested that pancreatic cancer patients with factors that may decrease the likelihood of resectability, such as elevated serum CA19-9 levels, should undergo staging laparoscopy.^6^ A number of studies have investigated potential preoperative predictive factors including tumor marker levels and tumor size that may determine actual resectability status of pancreatic cancer. It would therefore be helpful to have a synthesis of findings to see which parameters may or may not be helpful in the preoperative prediction of actual intraoperative resectability. This systematic review aims to identify parameters not part of radiologic resectability criteria, including clinical and laboratory markers, that may independently predict intraoperative resectability status for patients deemed to have potentially resectable pancreatic cancer.

## Methods

### Search Strategy

Literature search was done using the following databases: PubMed (MEDLINE), Scopus, Web of Science, and Cochrane Library. The following search string was used: (pancreatic cancer OR pancreatic malignancy OR pancreatic neoplasm OR pancreatic tumor OR pancreatic carcinoma OR pancreatic adenocarcinoma OR pancreatic ductal adenocarcinoma OR PDAC) AND (predictors OR prediction OR predicting OR indicators OR indications OR determinants OR markers OR clinical characteristics OR clinical profile OR clinicodemographic OR factors OR biological) AND (resectability OR resectable OR unresectability OR unresectable).

### Inclusion and Exclusion Criteria

The process of study selection for the systematic review adhered to the Preferred Reporting Items for Systematic Reviews and Meta-analyses (PRISMA) guidelines.^7,8^ After the initial search across databases, duplicate articles were removed first. The subsequent study inclusion criteria were set as follows: 1) studied population being patients deemed to have potentially resectable pancreatic cancer, 2) studied population subjected to attempted resection or staging laparoscopy to assess resectability, 3) availability of comparisons of preoperative data between subjects with resectable and unresectable pancreatic cancer, 4) study with analysis of factors that independently predict unresectable pancreatic cancer on exploration. Exclusion criteria were set as follows: 1) study done before the year 2011, 2) study mainly assessing accuracy of imaging modalities in determining resectability, 3) study exclusively assessing resectability after neoadjuvant treatment. Initial screening of articles was carried out by evaluating titles and abstracts, while final article selection was determined based on full-text assessment. The references of the final set of included studies were further assessed to determine any additional study that may be included in the systematic review based on the aforementioned inclusion and exclusion criteria.

### Quality Assessment

Quality assessment of the included studies was conducted using the Newcastle-Ottawa Scale.^9^

### Data Extraction and Analysis

The following data were extracted from the included studies: authors, year of publication, country of origin, study design, source of patient selection, inclusion and exclusion criteria for study subjects, number of study population, preoperative methods of resectability determination, use of staging laparoscopy, actual determination of resectability intraoperatively, predictive factors for resectability analyzed, and main findings. The main results in the included studies were then collated to identify which parameters have been found to be useful in the preoperative determination of actual surgical resectability by independently predicting intraoperative resectability status. Potential research gaps were also identified.

For predictive parameters investigated by at least two studies but with discordant findings, meta-analyses using the random effects model were done, which included studies that reported result data in a manner similar enough to allow pooling. Parameters with binary values were pooled to determine odds ratios (OR) with 95% confidence interval (CI), while those with continuous values were pooled to get weighted mean differences (WMD) with 95% CI. In the event of significant heterogeneity with I^2^ >50%, subgroup meta-analyses were done based on the following attributes: study design, region or country, inclusion of borderline resectable pancreatic cancer patients, inclusion of patients with any neoadjuvant treatment, and criteria in assessing intraoperative resectability. Studies with insufficient provided data that cannot be derived were excluded from meta-analyses. Funnel plots were generated and Egger’s regression tests were done for the analyses having at least 10 pooled studies to assess risk of publication bias.^10^ Sensitivity analyses were done by running the meta-analyses using the fixed effects model. For parameters whose studies cannot be pooled in meta-analyses, narrative synthesis was done.

### Conduction of the Systematic Review

Two reviewers independently performed the literature search, selection of studies for inclusion in the review, study quality assessment, extraction of data, and analysis of data. Any discrepancies encountered throughout the process were resolved via consultation and discussion with a third reviewer. Zotero was used as the reference manager in the conduction of the review. For meta-analyses, the MetaXL add-in for Microsoft Excel was used for the calculations and generation of forest plots.^11^ For generation of funnel plots and performance of Egger’s regression tests, JASP was used.^12^

## Results

### Overview

A total of 8959 articles were identified in the initial database search, of which 3171 were duplicates. The remaining 5788 articles were evaluated for eligibility using the predetermined inclusion and exclusion criteria, of which 14 were deemed eligible. On assessing the 564 total references across these 14 studies, an additional 4 more studies were found to satisfy the inclusion and exclusion criteria. Hence, a total of 18 studies were finally included in the analysis. Figure 1 shows the corresponding PRISMA diagram for the systematic review.

**Figure 1.**
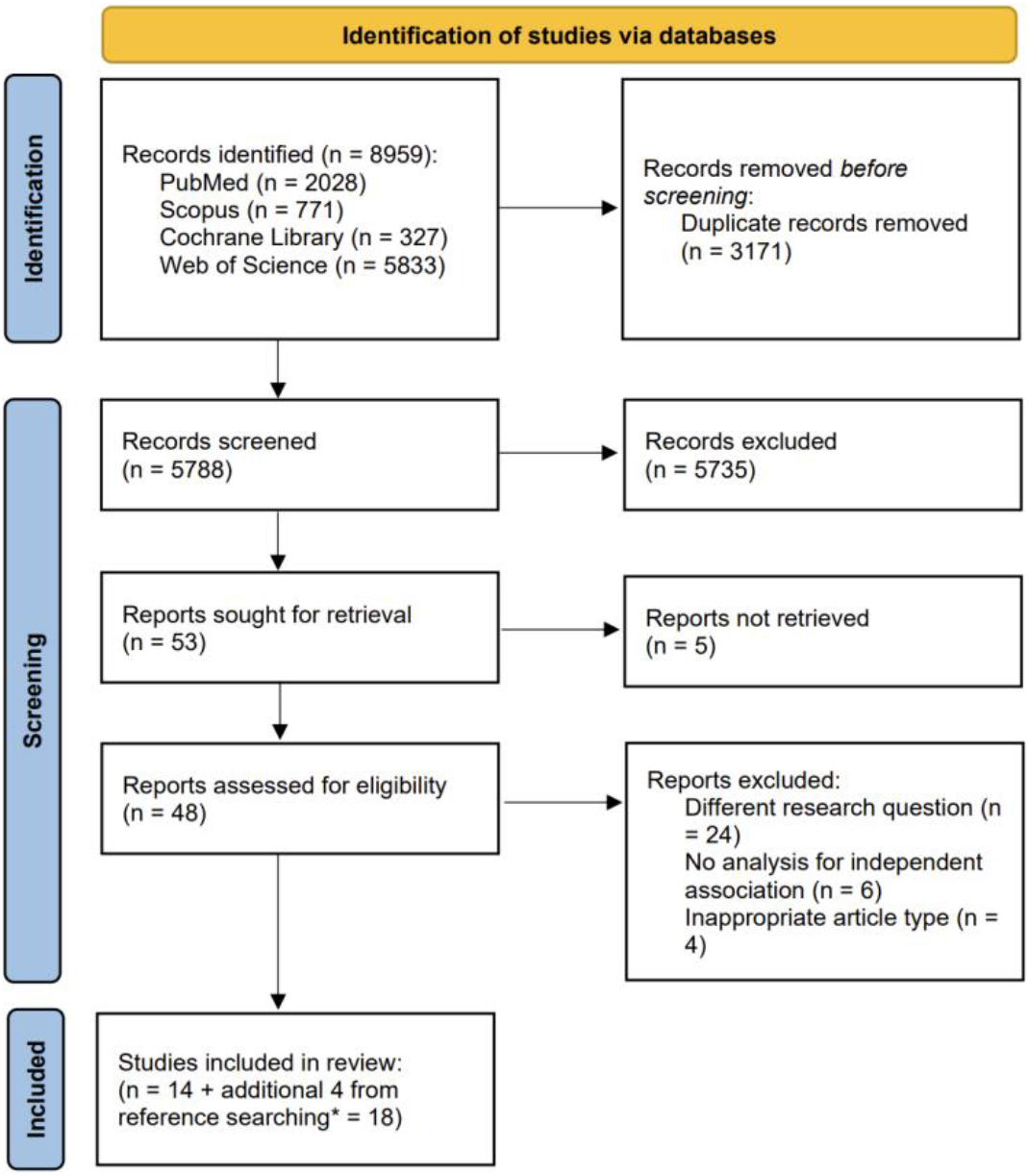
PRISMA Flow Diagram

### Quality Assessment

Assessment of the quality of studies included in the analysis was conducted in compliance with the Newcastle-Ottawa Quality Assessment Scale. In the evaluation of subjects included in the cohort, all studies are representative of the exposed cohort with the non-exposed cohort being drawn from the same community as the exposed group. Proper documentation of the cases were described using medical records with the outcome of interest of intraoperative resectability, by its nature, definitely not present at the start of the study. In terms of the comparability among cohorts in the studies, all studies controlled for potential confounding factors for every investigated parameter. The outcome measured was tied to the results as indicated in medical records. All included studies garnered 8 points each using the scale.

### Characteristics of the Included Studies

A total of 9,869 patients overall were analyzed in the 18 included studies, of which 10 were from East Asia, 5 were from Europe, and 3 were from the Americas. Three studies were prospective cohorts while the rest were retrospective analyses. All but three studies^22,24,30^ were conducted in single-center settings. All studies examined pancreatic cancer patients subjected to surgical exploration or staging laparoscopy for potentially resectable disease, with 11 studies^14,16,17,20,22,23,25–29^ specifying the inclusion of borderline resectable cases and 9 studies^15,17,19,20,23,25,26,28,29^ having patients with neoadjuvant treatment. With respect to outcome, 10 studies^14,16,17,19,20,23–25,29,30^ only looked into the detection of occult distant metastases on surgical exploration precluding curative resection, whereas 8 studies^13,15,18,21,22,26–28^ also included locally advanced disease encountered intraoperatively as a reason for unresectability. Table 1 summarizes the key attributes of the studies included in the systematic review.

**Table 1.** Summary of Studies Included in the Systematic Review.

| Study | Year | Country | Study Design | Patient Selection | Patient Number | Outcomes Assessed |
| --- | --- | --- | --- | --- | --- | --- |
| Chiang et al<sup>13</sup> | 2012 | Taiwan | Retrospective cohort | All studies included | 688 | Actual resectability |
| de Jesus et al<sup>14</sup> | 2018 | Brazil | Retrospective cohort | patients with potentially resectable pancreatic cancer who underwent curative-intent surgery or staging laparoscopy, with preoperative parameters assessed for independent association with actual intraoperative resectability status | 63 | (curative resection versus distant metastases and/or locally advanced disease at surgical exploration) |
| Eyff et al<sup>15</sup> | 2018 | Brazil | Retrospective cohort |  | 84 |  |
| Ge et al<sup>16</sup> | 2021 | China | Retrospective cohort |  | 273 |  |
| Gemenetzis et al<sup>17</sup> | 2018 | USA | Retrospective cohort |  | 468 |  |
| Hartwig et al<sup>18</sup> | 2013 | Germany | Prospective cohort |  | 1,543 |  |
| Hata et al<sup>19</sup> | 2021 | Japan | Retrospective cohort |  | 142 |  |
| Lee et al<sup>20</sup> | 2020 | South Korea | Retrospective cohort |  | 311 |  |
| Liu et al<sup>21</sup> | 2018 | China | Retrospective cohort |  | 235 |  |
| Maulat et al<sup>22</sup> | 2020 | France | Prospective cohort |  | 515 |  |
| Murakami et al<sup>23</sup> | 2024 | Japan | Retrospective cohort |  | 279 |  |
| Sahlstrom et al<sup>24</sup> | 2022 | Sweden | Retrospective cohort |  | 1,773 |  |
| Sakaguchi et al<sup>25</sup> | 2022 | Japan | Retrospective cohort |  | 405 |  |
| Sanjeevi et al<sup>26</sup> | 2015 | Sweden | Retrospective cohort |  | 349 |  |
| Satoi et al<sup>27</sup> | 2011 | Japan | Retrospective cohort |  | 94 |  |
| Shimizu et al<sup>28</sup> | 2021 | Japan | Retrospective cohort |  | 239 |  |
| Takadate et al<sup>29</sup> | 2021 | Japan | Retrospective cohort |  | 146 |  |
| Walma et al<sup>30</sup> | 2024 | Netherlands | Retrospective cohort |  | 2,262 |  |

### Factors Predictive of Intraoperative Resectability

Table 2 lists the parameters shown in at least one study to be independently associated with intraoperative resectability of pancreatic cancer on surgical exploration with initial curative intent. Elevated serum CA19-9 level was consistently reported as a predictive factor for intraoperative unresectability; however, different cut-offs were determined across studies, ranging from 35 to 1000 U/mL, above which unresectability may be anticipated.^13,16–21,23–25,27–29^ Larger tumor size, greater than 22-40 mm, was also frequently found to be an independent risk factor for actual pancreatic cancer unresectability.^13,14,17,21–23,25–28,30^ There were 3 studies that looked at serum aminotransferases: aspartate aminotransferase (AST) in all 3 studies^13,16,21^ and alanine aminotransferase (ALT) in 2 of them,^16,21^ with 2 studies finding them to be independently associated with intraoperative resectability status. Other parameters were less consistently found to be predictive of intraoperative resectability, including age, sex, abdominal pain, weight loss, body mass index (BMI), performance status, tumor location, and interval between imaging and surgery. The parameters of smoking status, neoadjuvant treatment, alcohol intake, hemoglobin level, neutrophil-to-lymphocyte ratio, platelet-to-lymphocyte ratio, and biliary drainage all had at least 2 studies unanimously showing their lack of independent association with intraoperative pancreatic cancer resectability. The presence of regional lymph node enlargement or suspicious lymph nodes on imaging was found to be associated with the occurrence of occult metastases and futile surgery.^16,20^ Furthermore, the presence of indeterminate lesions on imaging, such as in the liver or peritoneum, was associated with the presence of occult metastatic disease.^17,30^

**Table 2.** Identified Parameters for Synthesis.

| Type of Parameter | Parameter Assessed in at Least 2 Studies with Discordant Findings Regarding Predictive Value for Intraoperative Resectability | Number of Studies Finding the Parameter to be Independently Predictive of Intraoperative Resectability Status | Total Number of Studies Assessing Predictive Value of the Parameter |
| --- | --- | --- | --- |
| Clinical | Abdominal Pain | 2 | 5 |
|  | Performance Status | 1 | 3 |
|  | Weight Loss | 2 | 6 |
|  | Body Mass Index | 1 | 8 |
|  | Age | 2 | 15 |
|  | Sex | 1 | 15 |
| Laboratory | Serum CA 19-9 Level | 13 | 16 |
|  | Serum Transaminases | 2 | 3 |
|  | Serum CA 125 Level | 1 | 2 |
| Radiologic | Tumor Size | 11 | 15 |
|  | Borderline Resectability | 2 | 5 |
|  | Interval between Imaging and Surgery | 1 | 4 |
|  | Tumor Location | 4 | 13 |

There were no notable discrepancies in the findings between prospective and retrospective studies. Likewise, there also do not seem to be major differences in the findings from studies originating from developed countries compared to those from developing countries. Only the studies from China and Taiwan, which also specified the exclusion of patients with neoadjuvant treatment, explored the factor of serum transaminases. Furthermore, among these, only those that just used the presence of occult metastatic disease on exploration as outcome detected a predictive value of serum transaminases in determining actual resectability of pancreatic cancer.

### Meta-analysis

The parameters of abdominal pain, performance status, weight loss, BMI, age, sex, serum transaminases, preoperative borderline resectable status, and tumor location were subjected to meta-analyses (Figure 2). On the other hand, the findings on the parameters of serum CA19-9, serum CA125, tumor size, and interval between imaging and surgery could not be pooled due to differences in the data presentation across studies.

**Figure 2.**
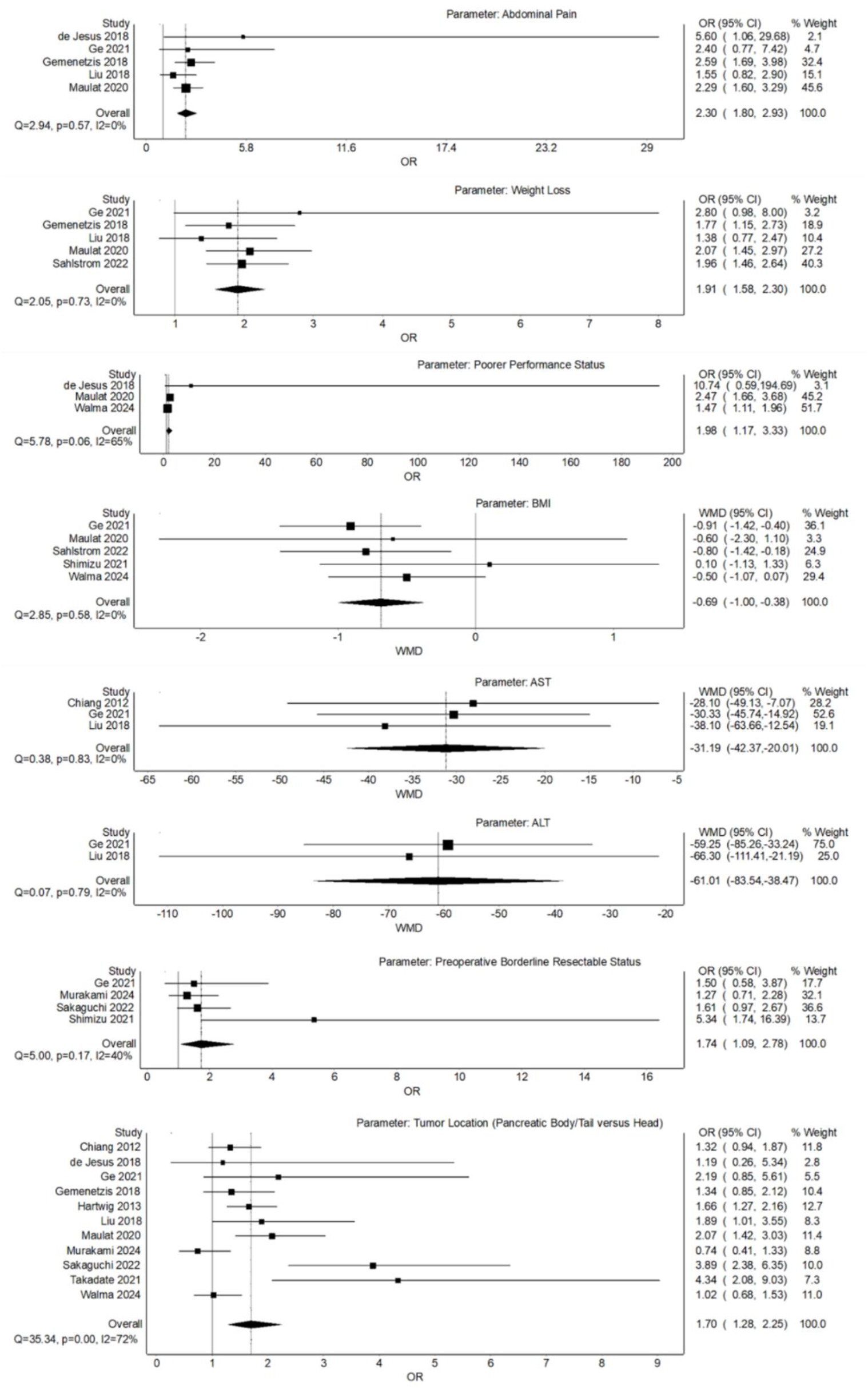
Forest Plot Analyses for the Parameters of Abdominal Pain, Weight Loss, Poorer Performance Status, BMI, AST, ALT, Preoperative Borderline Resectable Status, and Tumor Location.

For parameters with binary variables, the presence of abdominal pain (OR 2.30 [95% CI 1.80-2.93]), presence of weight loss (OR 1.91 [95% CI 1.58-2.30]), and preoperative borderline resectable status (OR 1.74 [95% CI 1.09-2.78]) were found to be significant risk factors for intraoperative unresectability. Performance status and tumor location were shown to be significant predictors of intraoperative resectability, but their analyses had significant heterogeneity (I^2^ = 65% and I^2^ = 72%, respectively). On subgroup analysis (Figure 3) that only included the two prospective cohort studies which resulted in insignificant heterogeneity, tumor location still predicted intraoperative resectability status (OR 1.78 [95% CI 1.44-2.22]). On the other hand, for the parameter of performance status, subgroup analysis including only the two retrospective studies, which addressed heterogeneity, revealed that it no longer independently predicted intraoperative resectability (OR 2.31 [95% CI 0.45-11.75]).

**Figure 3.**
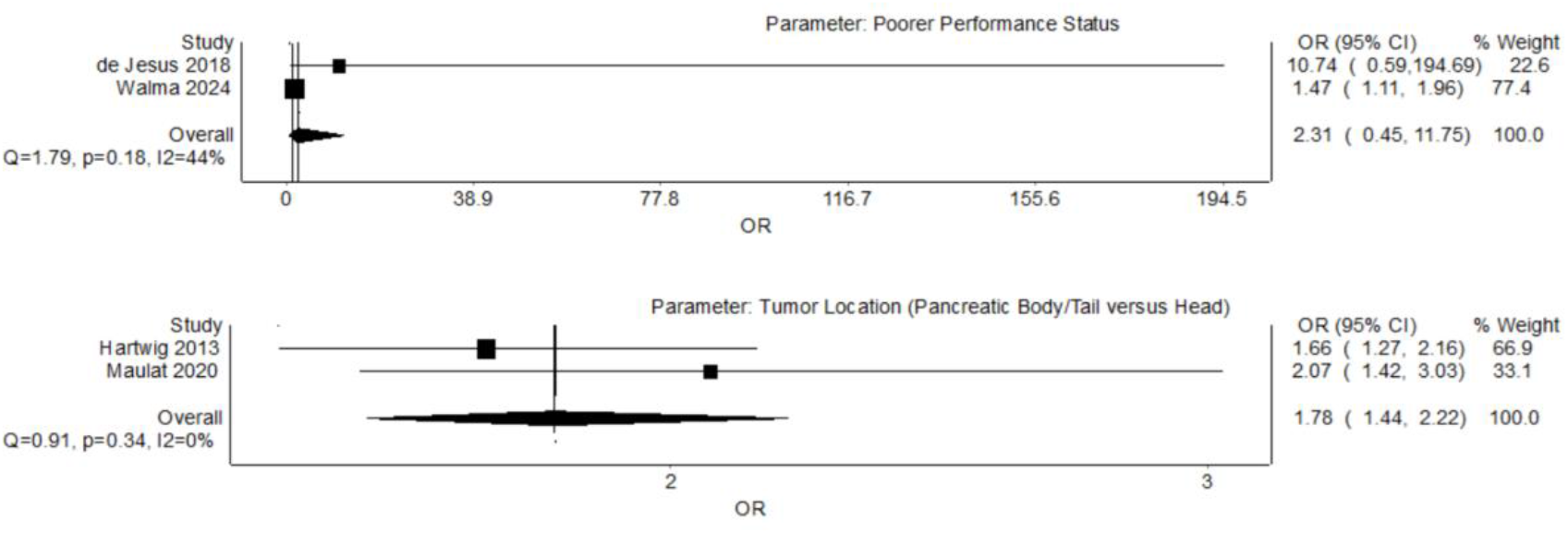
Subgroup Analyses Addressing Heterogeneity for the Parameters of Performance Status and Tumor Location.

For continuous variables, WMDs were used to measure if there were substantial differences among the studies selected. There was significantly lower mean BMI in the intraoperatively unresectable group than in the resectable group (WMD -0.69 [95% CI -1.00 to -0.38]). The mean levels of serum aminotransferases were also found to be significantly lower in the intraoperatively unresectable group than in the resectable group (WMD -31.19 [95% CI -42.37 to -20.01] for AST and WMD -61.01 [95% CI -83.54 to -38.47] for ALT). All continuous variables mentioned demonstrated statistically significant differences favouring lower values in the unresectable pancreatic cancer cohort.

The parameters of age (WMD -0.48 [95% CI -0.67 to 1.62]) and sex (OR 0.99 [95% CI 0.88-1.11]) were not significantly different between actual resectable and unresectable groups. Funnel plot analysis for both parameters (Appendix) indicate no significant asymmetry suggesting a low risk for publication bias, further supporting that these factors are not associated with pancreatic cancer resectability. Egger’s regression tests for the parameters of age (p = 0.088) and sex (p = 0.907) were also not significant (Appendix).

## Discussion

Surgical resection remains the only potential curative treatment for pancreatic cancer.^1,2,31,32^ Assessment of resectability has widely relied on determination of distant metastases and vascular invasion via imaging studies.^3,4,33,34^ However, accurate determination of the resectability of pancreatic cancer remains challenging despite advancements in radiology. For instance, accurate detection of liver and peritoneal metastases and determination of vascular invasion via multidetector computed tomography (MDCT) are still major issues.^35^ Around 5% to 15% of patients predicted to have resectable disease based on CT scan have been found at laparoscopy to have unresectable lesions.^4^ In addition, a similar percentage of patients that appear to have resectable disease by CT criteria have been found to have unresectable disease at exploratory surgery.^4^ Redefining resectability criteria beyond imaging-based morphological criteria has thus been proposed.^36^ In this study, we aimed to determine parameters that may predict actual intraoperative resectability outside resectability criteria which are mainly determined by preoperative radiologic imaging.

Serum CA19-9 levels have been shown to be a prognostic factor in pancreatic cancer, even among patients with jaundice from cholestasis.^37^ Additionally, in this systematic review, elevated serum CA19-9 level has been consistently found to be independently predictive of intraoperative unresectability of pancreatic cancer. This is in line with an international consensus by the IAP which considered a serum CA19-9 level of greater than 500 U/mL as a biological factor that potentially heralds metastases in potentially resectable disease.^5^ However, across studies investigating the utility of serum CA19-9 in predicting actual pancreatic cancer resectability, there is a wide range of cut-off levels that were used (35 to 1000 U/mL). It would thus appear that the predictive value of serum CA19-9 with respect to pancreatic cancer resectability may be more useful in determining actual resectable disease given a normal or low result. Furthermore, there may also be limitations in using serum CA19-9 as it has been reported that approximately 10% of Caucasians lack Lewis antigen A that is necessary to express CA19-9.^24,38^

Tumor size, which is not part of resectability criteria, may actually help predict intraoperative pancreatic cancer resectability. This systematic review found that high tumor size, greater than the range of 22-40 mm, has been shown to be independently associated with a high likelihood of occult metastasis in several studies. Measurement of tumor size was usually performed using CT although some studies opted for endoscopic ultrasound especially for tumors measuring 30 mm or smaller, as it can provide better resolution.^23^ With respect to tumor location, this meta-analysis revealed that pancreatic cancer located in the pancreatic body or tail increased the risk of intraoperative unresectability, even after subgroup analysis that addressed heterogeneity. This is consistent with the epidemiology that patients with pancreatic body or tail cancers are usually diagnosed later and have worse prognosis than those with pancreatic head cancer.^39,40^ These findings support the idea of revisiting resectability criteria to incorporate tumor size and location, or at least taking them into consideration before proceeding with curative-intent surgery among patients with potentially resectable pancreatic cancer.

A previous study suggested that clinical signs and symptoms may give clues regarding the resectability of pancreatic cancer.^41^ Abdominal pain was more frequently present in patients with unresectable pancreatic cancer while jaundice was more frequently present in patients with resectable disease. On the other hand, there were no significant differences between the resectable and unresectable pancreatic cancer groups in terms of the parameters of age, sex, and presence of weight loss. In support of these findings, meta-analysis in this study showed that the presence of abdominal pain is indeed a significant predictor of intraoperative unresectability. This is consistent with the pathophysiology of abdominal pain in pancreatic cancer in that pain would be expected in more advanced disease enough to cause invasion of the celiac or superior mesenteric arterial plexus resulting in neuropathy.^4,42,43^ Whereas age and sex were also not found to be significant predictors of intraoperative resectability in this meta-analysis, weight loss was found to be predictive of intraoperative unresectability, in contrast to the findings of the aforementioned study. In relation to this, a significantly lower BMI was also seen in those with actual unresectable pancreatic cancer. These findings are more consistent with what would be anticipated in unresectable disease since the presence of malnutrition, sarcopenia, and cachexia are common in advanced tumor stages of pancreatic cancer.^44,45^

The association of poorer performance status with intraoperative unresectability in this meta-analysis appears consistent with an international consensus that a performance status of 2 or more is a conditional factor for borderline resectability.^5^ However, this parameter seems to be a less robust indicator given the significant heterogeneity among the included studies. A more peculiar finding in this meta-analysis is that serum transaminase values were noted to be much lower in the unresectable group than in those who successfully underwent curative resection. Intuitively, patients with lower serum transaminases would be expected to have a less severe condition which should indicate more resectable disease. Nonetheless, a possible explanation is that patients with lower serum transaminases, particularly those without distant metastases, may have fewer symptoms and therefore get diagnosed later in the course of pancreatic cancer due to more delayed presentation.^16,46^

Overall, the findings in this study support the notion that pancreatic cancer patients having symptoms suggestive of more advanced disease may be expected to have actual unresectable tumors despite being deemed resectable by preoperative resectability criteria. These findings may serve as a guide in selecting patients for staging laparoscopy rather than proceeding with outright curative-intent surgery in order to prevent unnecessary laparotomies. Given that there are multiple factors associated with unresectability, it is still important to approach each case in coordination with a multidisciplinary team in order to optimize patient outcomes.^33^

A limitation of this study is that the parameters were considered individually. Future clinical studies may therefore investigate how combinations of these preoperative parameters affect the prediction of intraoperative resectability and attempt to develop a unified scoring system to aid clinical decision-making. Furthermore, studies are also needed from geographic areas that this systematic review has found to be unrepresented, particularly in Southeast Asia, where the evaluation of patients with suspected pancreatic cancer may be confounded by mimics like tuberculosis.^47–49^

## Conclusion

Elevated serum CA19-9 level, larger tumor size, and presence of indeterminate lesions or enlarged lymph nodes on imaging seem to reliably predict actual intraoperative unresectable disease among pancreatic cancer patients considered for curative-intent surgery. The presence of abdominal pain, weight loss, preoperative borderline resectable status, and tumor location in the pancreatic body or tail also appear to increase risk for unresectable disease on surgical exploration, whereas age, sex, smoking status, neoadjuvant treatment, interval between imaging and surgery, alcohol intake, hemoglobin level, neutrophil-to-lymphocyte ratio, platelet-to-lymphocyte ratio, and biliary drainage do not appear to have predictive value. Poorer performance status appears to have less robust evidence of independent association with actual unresectable disease. Lower serum aminotransferase level is a potential independent risk factor for actual unresectability; however, further studies are needed to confirm and elucidate the underlying mechanism, such as via correlation with time intervals from symptom onset to first consultation. Research on this topic is needed from other global regions like Southeast Asia.

## Data Availability

All data produced in the present study are available upon reasonable request to the authors.

## Appendix

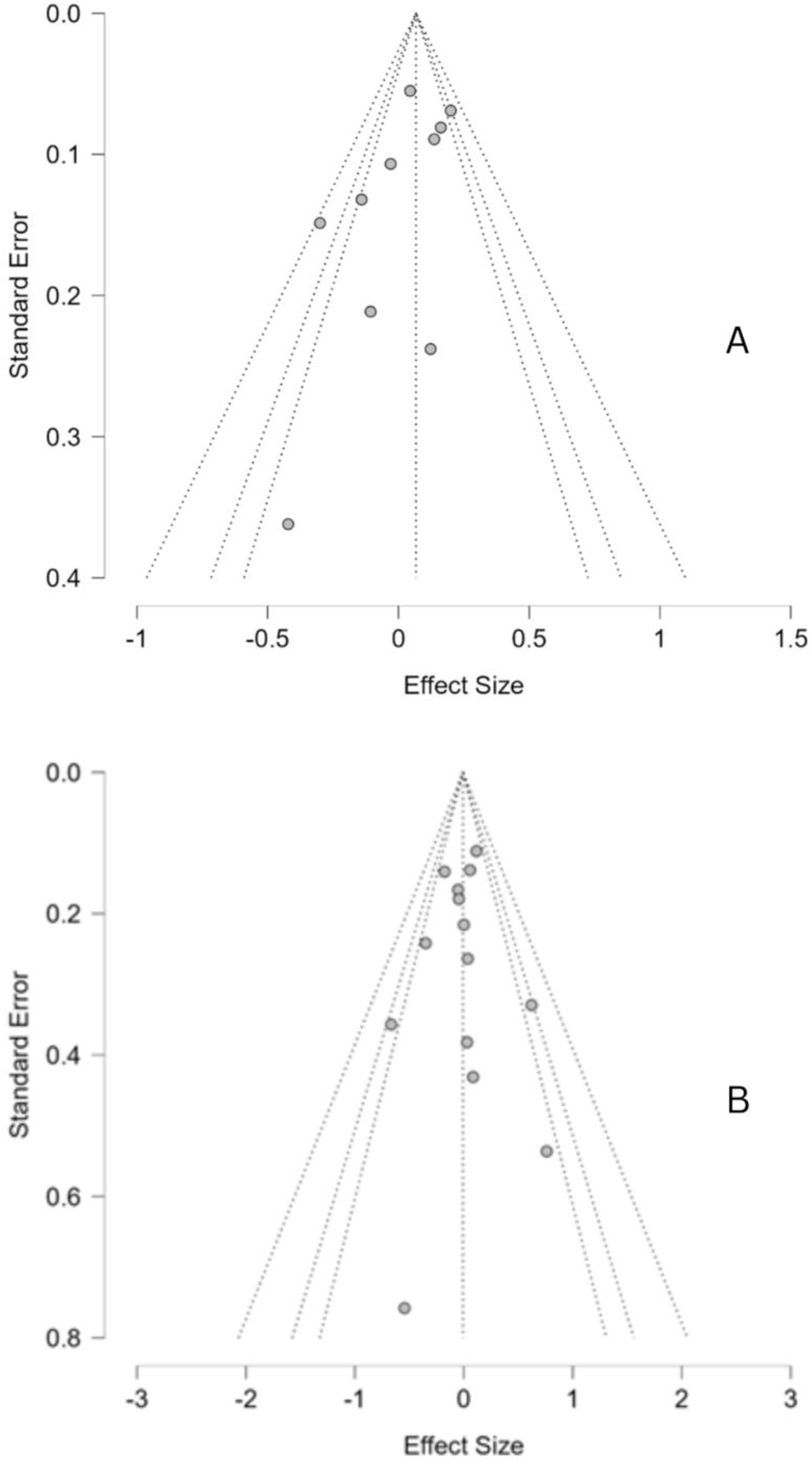

Funnel Plots for the Parameters of Age (A) and Sex (B).

Weighted Regression (Egger’s) Test for Funnel Plot Asymmetry (For Parameter of Age)

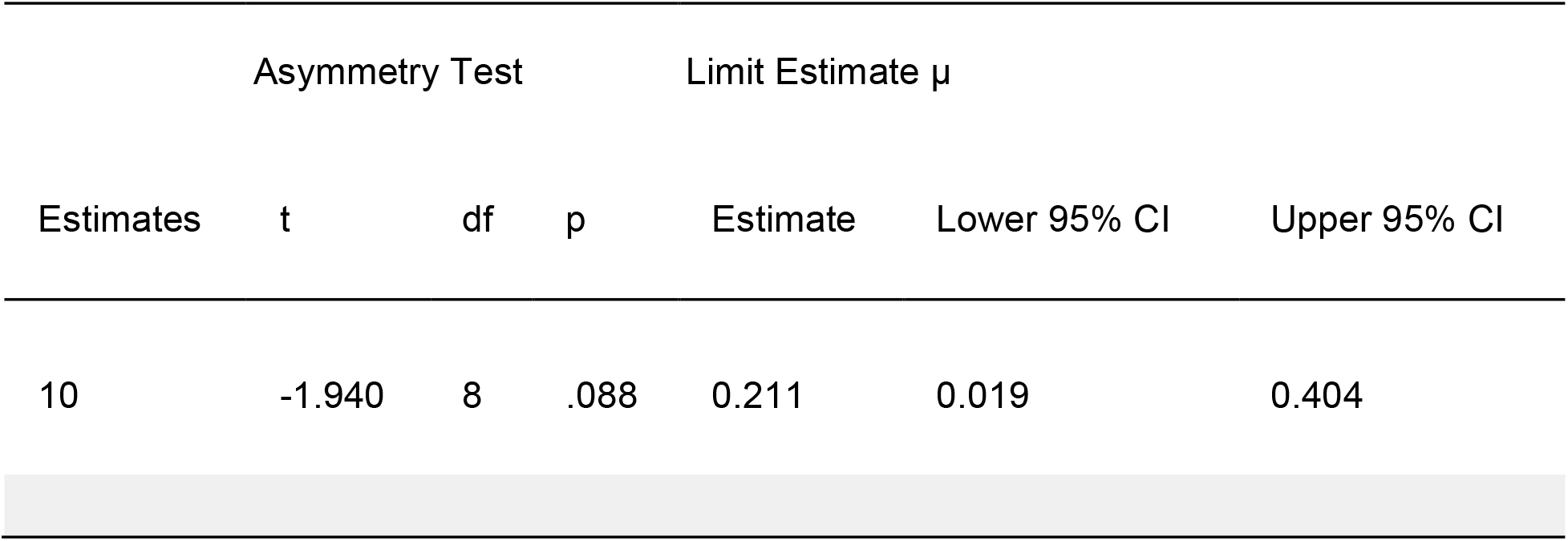

Weighted Regression (Egger’s) Test for Funnel Plot Asymmetry (For Parameter of Sex)

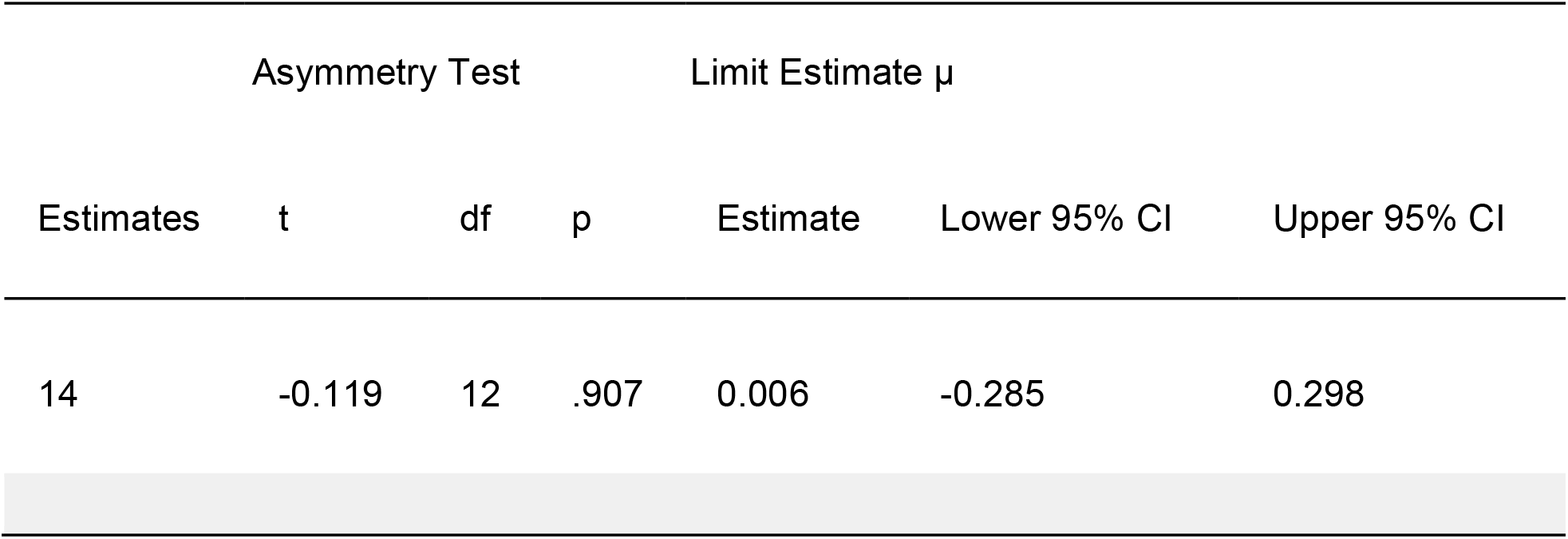

